# Psychomotor retardation and risk of Parkinson’s disease in unipolar depression: a retrospective cohort study

**DOI:** 10.64898/2026.04.26.26351763

**Authors:** Hamilton Morrin, James B. Badenoch, Ella Burchill, Aurore Fayosse, Archana Singh-Manoux, Paul Shotbolt, Michael S. Zandi, Anthony S. David, Glyn Lewis, Jonathan P. Rogers

## Abstract

**Background:** Depression is associated with an increased risk of subsequent Parkinson’s disease. Neuroimaging studies suggest a neurobiological overlap in mechanisms underlying Parkinson’s disease and psychomotor retardation in depression. Our aim was to investigate whether, among individuals with depression, the presence of psychomotor retardation was associated with the development of subsequent Parkinson’s disease.

**Methods:** In a retrospective cohort study, electronic healthcare records from individuals diagnosed with depression at age 40 or over in a large mental health service in London, UK were examined for the presence of psychomotor retardation. Linkage to general hospital records was used to ascertain diagnoses of Parkinson’s disease between 2007 and 2023. Cox regression was used to compare the hazard of Parkinson’s disease in individuals with depression with and without psychomotor retardation.

**Results:** Among 6327 patients with depression, 2402 (38.0%) had psychomotor retardation. The adjusted hazard ratio for development of Parkinson’s in those with psychomotor retardation was 1.43 (95% CI 1.02 - 2.01, *p* = 0.04). Secondary analyses demonstrated a significant difference in psychomotor retardation incidence at least 10 years before Parkinson’s diagnosis.

**Conclusions:** Psychomotor retardation in later-life depression is associated with increased risk of subsequent Parkinson’s diagnosis over an extended period of time, suggesting that the relationship cannot solely be explained by misdiagnosis. Psychomotor retardation may therefore serve as a marker of prodromal Parkinson’s disease.

**Highlights:**

- Psychomotor retardation was associated with later Parkinson’s disease.
- Psychomotor retardation may present >10 years prior to Parkinson’s diagnosis.
- Depression with psychomotor retardation may be a prodrome for Parkinson’s disease.

## 1. Introduction

Depression is a leading cause of disability globally, affecting 5% of adults in global estimates (World Health Organisation, 2025). Previous studies have shown a strong relationship between depression and Parkinson’s disease (PD). Clinically significant depressive symptoms are present in around 35% of cases of PD (range 2.7 - 90%) (Reijnders et al., 2008). In recent years, it has become clear that the relationship between Parkinson’s disease and depression may in fact be bidirectional, with a Swedish nationwide cohort study finding an odds ratio for PD development in individuals with depression of 3.2 (95% CI 2.5 - 4.1) within the first year of depression, and 1.5 (95% CI 1.1 - 2.0) after 15 to 25 years (Gustafsson et al., 2015). As a result, depression has been proposed as a prodromal symptom and risk factor for PD. However, to date no studies to our knowledge have focused on whether specific depressive psychopathology increases risk of PD over the long-term.

Psychomotor retardation (or psychomotor slowing) is a psychopathological feature of depression with considerable implications for disease severity and response to treatment. It is defined as a slowing down of thought and speech, with impaired cognitive function and reduced physical movement (Buyukdura et al., 2011). Psychomotor retardation is often associated with more severe depressive episodes, including episodes of psychotic depression (Janzing et al., 2020). It is also an independent predictor of response to electroconvulsive therapy (Heijnen et al., 2019; van Diermen et al., 2020), suggesting a distinct pathophysiology.

There are phenomenological similarities between depression with psychomotor retardation and Parkinson’s disease. Both can feature slowed movement, poverty of emotional expression and quiet speech (Leong et al., 2026). Subtle motor abnormalities, such as early gait changes, have been identified as potentially helpful in identifying those clinically at risk of developing PD (Del Din et al., 2019). Findings from structural and functional neuroimaging studies suggest that deficits in the basal ganglia, prefrontal cortex, and striatal dopaminergic pathways may play a role in the neurobiological underpinnings of psychomotor retardation, pathways also implicated in PD (Bennabi et al., 2013; Buyukdura et al., 2011; Leong et al., 2026). More recent studies have identified a role for the glymphatic system in pathophysiology of both depressive psychomotor retardation (Liang et al., 2025) and motor symptoms in PD (Rajai Firouzabadi et al., 2025). These findings raise the question as to whether psychomotor retardation may be an early marker of neurodegeneration associated with PD.

This study aimed to explore the association between psychomotor retardation in depression and subsequent risk of Parkinson’s disease in a retrospective cohort of individuals diagnosed with depression. We hypothesised that in individuals with depression, the presence of psychomotor retardation would be associated with a greater risk of incident PD diagnosis. By identifying whether psychomotor retardation is a predictor of future PD, this research may contribute to refining early detection strategies and improving long-term outcomes for individuals at risk of neurodegeneration.

## 2. Methods

### 2.1. Study design

A retrospective cohort study design using data recorded between 01/01/2007 and 31/03/2023 was used. Participants were identified using the Clinical Records Interactive Search (CRIS) system, which consists of anonymised electronic healthcare records from South London and Maudsley NHS Foundation Trust, a large provider of secondary mental health services in London, UK (Perera et al., 2016).

Participants were included if a diagnosis of unipolar depression (defined as an ICD-10 diagnosis of F32 – Depressive episode or F33 – Recurrent depressive disorder) was made at age 40 years or older. Participants were also required to have at least one mention of psychomotor status following depression diagnosis. The index date for survival analysis was defined as the date on which exposure status was assigned: the earliest recorded psychomotor retardation classification after depression diagnosis for exposed individuals, and the earliest psychomotor-status record after depression diagnosis for individuals without any recorded psychomotor retardation. Participants were excluded if they had a diagnosis of Parkinson’s disease prior to depression diagnosis.

A full list of variable definitions is provided in Supplementary Table 1. The exposure (psychomotor retardation) was defined using a natural language processing (NLP) application, which used the free text of patients’ notes. The NLP application was validated in a sample of 50 positive hits and 50 negative hits by a psychiatrist with expertise in movement disorders (JPR) and showed a positive predictive value of 74% and a negative predictive value of 100%. Patients were classified by the application as having psychomotor retardation, psychomotor agitation or no psychomotor abnormalities. Patients with any psychomotor retardation following depression diagnosis were considered as exposed, while those with either psychomotor agitation or no psychomotor abnormalities coded after depression diagnosis (with no psychomotor retardation) were considered unexposed.

Since diagnoses of Parkinson’s disease are underestimated in routinely collected data (Muzerengi et al., 2017), we used three complementary methods to capture Parkinson’s diagnoses: (1) a diagnosis of Parkinson’s disease coded in CRIS with an ICD-10 code of G20 or F02.3; (2) a diagnosis of Parkinson’s disease in the free text of CRIS identified using the Medical Concept Annotation Tool (MedCAT), an NLP application (Kraljevic et al., 2021); and (3) a diagnosis of Parkinson’s in linked Hospital Episodes Statistics (HES) with an ICD-10 code of G20 or F02.3 (Jewell et al., 2020). No codes that had an overlap with drug-induced parkinsonism were included. The outcome was defined as the earliest diagnosis of Parkinson’s disease identified by any of these three methods.

Follow-up was conducted from index date until development of Parkinson’s disease, death or 31/03/2023, whichever was earliest.

The manuscript is written according to the STROBE guidelines (von Elm et al., 2008) and the checklist is present in Supplementary Table 2.

### 2.2. Covariates

Covariates chosen in the study were gender, antipsychotic medication use, major neurological disorders (multiple sclerosis, cerebrovascular accident, epilepsy, and transient ischaemic attack) and other major psychiatric disorders (ICD-10 F0–9, and F20-29). These variables were selected on the basis that they theoretically predated the exposure (psychomotor retardation) and may be confounders of any relationship to Parkinson’s disease. The exception was antipsychotic use, which we adjusted for in order to avoid our effect being mediated by prescribing patterns that might unmask Parkinson’s disease. Participant age was used as the timescale in order to handle an anticipated non-linear relationship between age and PD risk. Individuals with missing data for gender or age at date of censoring were removed from the dataset.

### 2.3. Sample size

The sample size was obtained by identifying all eligible individuals in the CRIS database.

### 2.4. Statistical analysis

Descriptive statistics were displayed by exposure status, using number of participants and percentages for categorical variables, and median and interquartile range for numeric variables. The primary analysis was a Cox proportional hazards regression adjusting for gender, White ethnicity, age at depression diagnosis, antipsychotic use, major neurological comorbidity and major psychiatric comorbidity. The regression model used age as the timescale in anticipation of PD risk varying in a non-linear way across age groups. Results were presented unadjusted and adjusted for covariates. A Kaplan-Meier curve was used for graphical representation.

Due to the potential for varying diagnostic patterns over calendar time, as a sensitivity analysis, Lexis expansion dividing calendar time into 2-year periods was conducted and the primary analysis repeated on the expanded data (Hong and Lewington, 2019). A further sensitivity analysis was conducted in which individuals whose first psychomotor exposure status was within 1 year preceding censor were excluded in order to reduce the potential for misdiagnosis of psychomotor retardation as Parkinson’s disease. Additional sensitivity analyses were conducted excluding patients using antipsychotics and using only PD diagnoses from HES records.

We conducted a secondary analysis to ascertain the longitudinal relationship between psychomotor retardation and PD, i.e. specifically to determine when differences in psychomotor retardation begin to emerge between individuals who do and do not go on to develop PD. To do this, we used a logistic mixed-effects model with a backwards timescale (Singh-Manoux et al., 2017), where exposures to psychomotor retardation were clustered within individuals. Time from exposure status to censor was fitted as both a linear and a quadratic term to account for non-linear relationships. Exposure status was regressed onto time, time-squared, Parkinson’s disease diagnosis, age at censor, gender, antipsychotic use, major neurological comorbidity and major psychiatric comorbidity. An interaction between time and Parkinson’s disease, and between time-squared and Parkinson’s disease was fitted.

Statistical analyses were conducted in *R* version 4.3.0 using RStudio 2023.06.0 and the survival version 3.5-7, survminer version 0.4.9, rstpm2 version 1.6.2, LexisPlotR version 0.4.0, lme4 version 1.1-34, lmerTest version 3.1-3, nlme version 2.1-162, and Epi version 2.58 packages. The statistical code is available in Supplementary Methods 1. The threshold for statistical significance was set to 0.05.

## 3. Results

The search identified 24,973 patients diagnosed with unipolar depression at age >40 years in CRIS. Of these, 6327 had at least one psychomotor exposure status recorded: 2402 were recorded as having psychomotor retardation and 3925 without psychomotor retardation. Participants are compared in terms of baseline characteristics in Table 1.

**Table 1:** Baseline demographic and clinical characteristics of exposure groups.

|  | Psychomotor retardation present (*N* = 2402) | Psychomotor retardation absent (*N* = 3925) |
| --- | --- | --- |
| Gender* |  |  |
| - Male, *n* (%) | 1040 (43.3) | 1759 (44.8) |
| - Female, *n* (%) | 1362 (56.7) | 2166 (55.2) |
| Age at first depression diagnosis, median (IQR) | 55.0 (46.1 - 68.0) | 53.0 (45.6 - 66.0) |
| Ethnicity |  |  |
| - White, *n* (%) | 1425 (59.4) | 2462 (62.7) |
| - Black / African / Caribbean / Black British, *n* (%) | 522 (21.7) | 533 (13.6) |
| - Asian / Asian British | 176 (7.3) | 210 (5.4) |
| - Mixed / multiple ethnic groups, *n* (%) | 23 (1.0) | 51 (1.3) |
| - Other ethnic groups, *n* (%) | 226 (9.4) | 426 (10.9) |
| - Not stated, *n* (%) | 30 (1.2) | 63 (1.6) |
| Major neurological comorbidity, *n* (%)<sup>†</sup> | 54 (2.2) | 146 (3.7) |
| Major psychiatric comorbidity, *n* (%)<sup>†</sup> | 155 (6.5) | 199 (5.1) |
| Antipsychotic use, *n* (%)<sup>†</sup> | 1407 (58.6) | 1490 (38.0) |
| ECT<sup>†</sup> | 240 (10.0) | 95 (2.4) |
| Deaths<sup>†</sup> | 736 (30.6) | 1072 (27.3) |
| Observation time (years), median (IQR) | 5.75 (2.50 - 10.4) | 5.44 (2.38 - 9.40) |
| Incident Parkinson's disease diagnosis, *n* (%) | 72 (3.0) | 67 (1.7) |
| Time from depression to Parkinson's disease diagnosis (years), median (IQR) | 3.58 (1.22 - 6.57) | 3.41 (1.02 - 8.50) |
| Age at censor, median (IQR)<br>* | 63.0 (55.6 - 74.7) | 61.5 (54.2 - 72.8) |

SD – standard deviation, IQR – interquartile range
\* No missing data, as only participants with complete data on these measures were eligible.
<sup>†</sup> In common with usual practice in electronic healthcare record research, we assumed that an absence of mention of these binary variables meant that they were not present.

Among those individuals with psychomotor retardation, 72 (3.0%) developed Parkinson’s disease, compared to 67 (1.7%) without psychomotor retardation. A Kaplan-Meier curve for the development of Parkinson’s disease is shown in Figure 1. In the unadjusted Cox proportional hazards regression, this corresponded to a hazard ratio of 1.55 (95% CI 1.11 – 2.16, *p* = 0.01). In the adjusted Cox proportional hazards regression, the hazard ratio was 1.43 (95% CI 1.02 - 2.01, *p* = 0.04).

**Figure 1:**
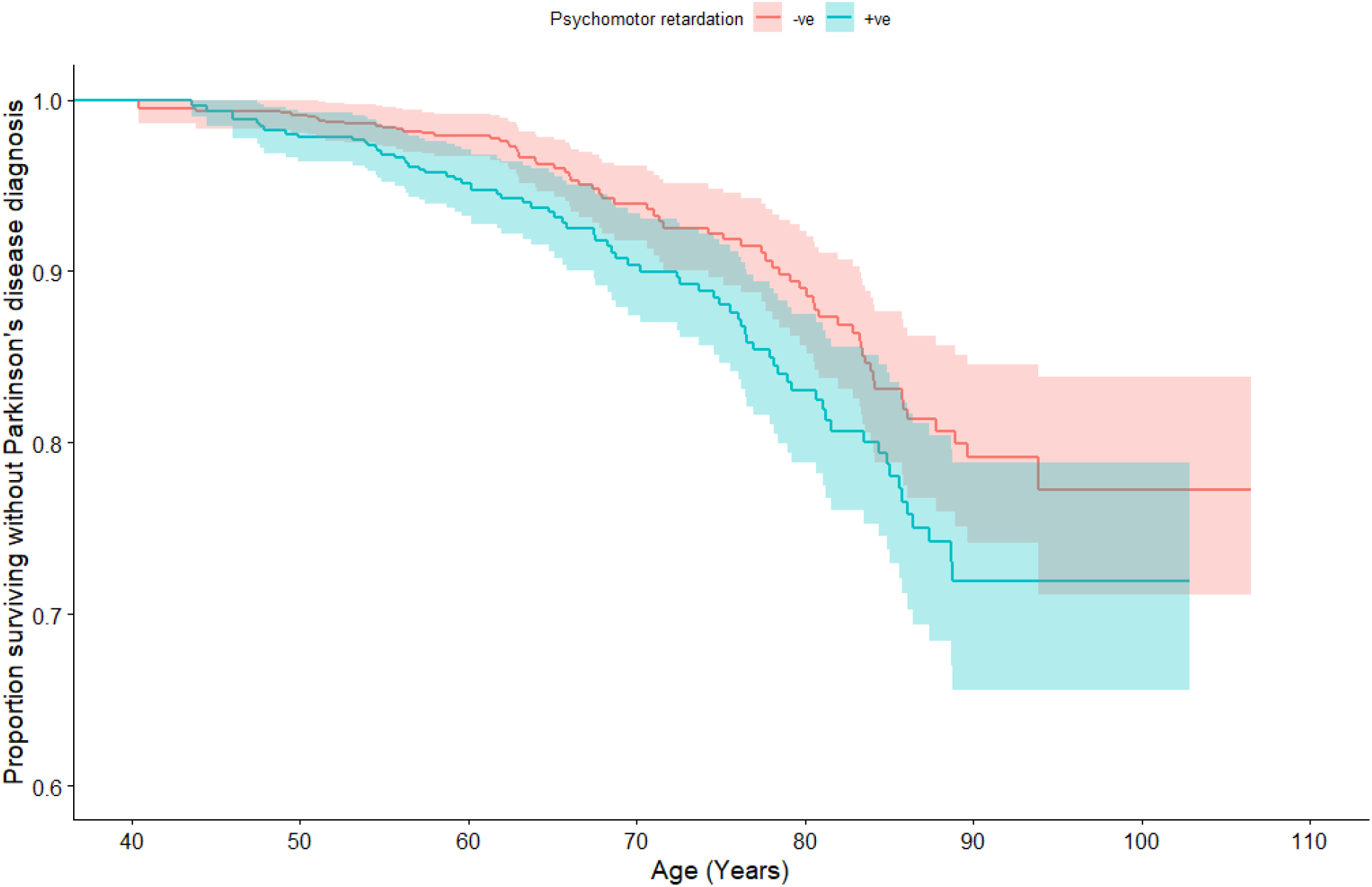
Kaplan-Meier curve for development of Parkinson’s disease in depressed individuals with and without psychomotor retardation

Results for the sensitivity analyses showed similar hazard ratios to the main analysis and are shown in Table 2.

**Table 2:** Results of sensitivity analyses.

| Sensitivity analysis | Adjusted hazard ratio for psychomotor retardation status (95% CI) | *p* |
| --- | --- | --- |
| Lexis expansion using calendar year | 1.39 (0.99 – 1.96) | 0.06 |
| Excluding individuals with a first exposure status within 1 year of censor | 1.37 (0.90 - 2.08) | 0.14 |
| Excluding patients using antipsychotic medications | 1.39 (0.80 - 2.34) | 0.24 |
| Using only HES diagnoses of Parkinson's disease | 1.29 (0.80 - 2.08) | 0.29 |

CI – confidence interval. HES – Hospital Episode Statistics.

In the backwards logistic mixed-effects model with a quadratic time interaction, there were significant effects observed for the linear component of time (p < 0.001) and quadratic component of time (p = 0.005), indicating that the probability of psychomotor retardation changed over time in a non-linear manner. Figure 2 is a LOESS-smoothed plot of observed psychomotor retardation over backwards time by Parkinson’s status and illustrates that the rates of psychomotor retardation are consistently and reliably higher in the Parkinson’s disease group at least 10 years before censor date.

**Figure 2:**
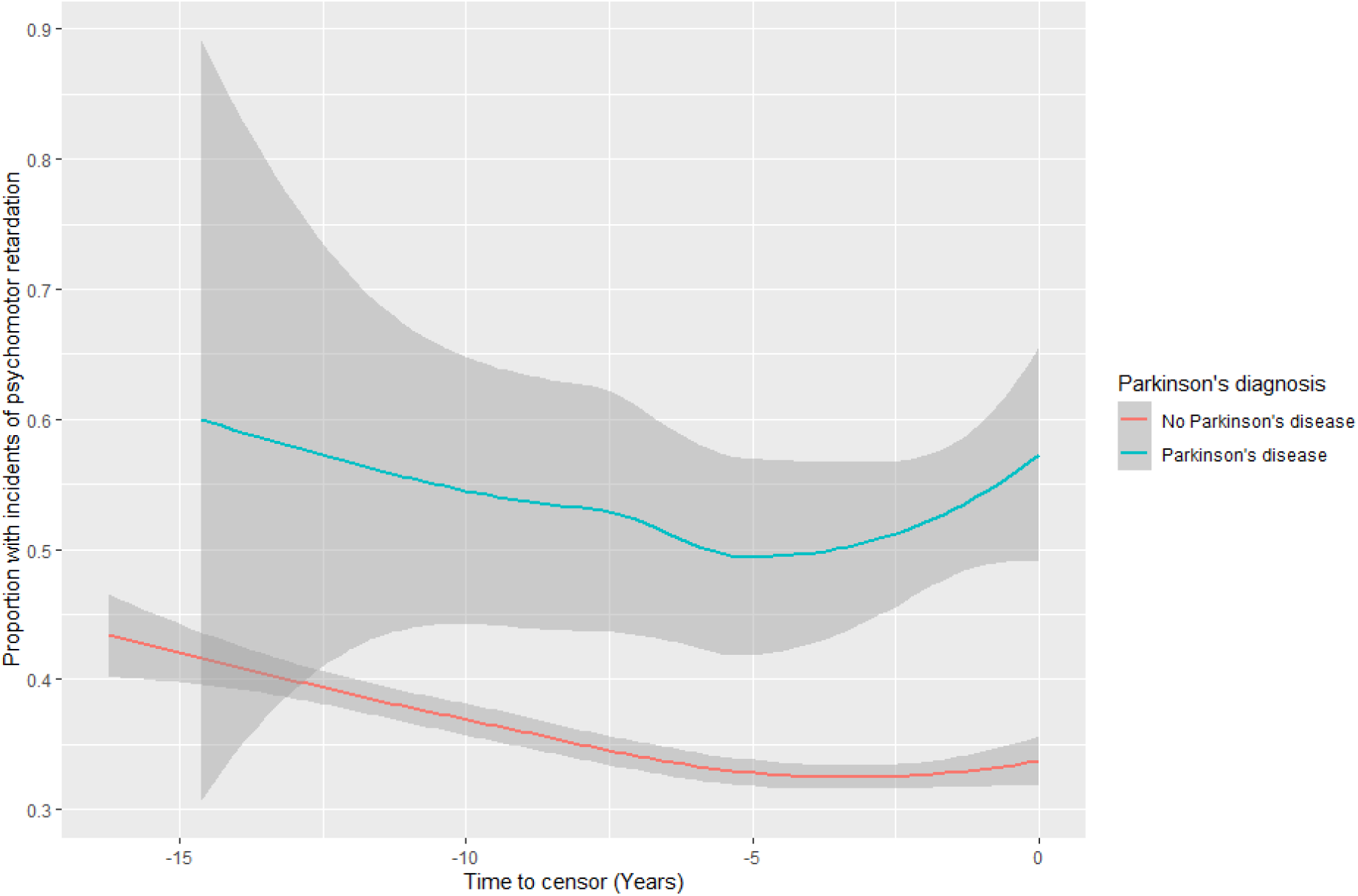
Graph demonstrating proportion of participants with psychomotor retardation prior to censor date

## 4. Discussion

To the authors’ knowledge, this retrospective cohort study is the first to explore the relationship between specific depressive psychopathology and development of Parkinson’s disease. Our findings suggest that psychomotor retardation in depression is associated with a higher subsequent risk of PD (HR = 1.43, 95% CI 1.02 - 2.01), and our secondary analysis suggests that this increased risk dates back to at least 10 years prior to PD diagnosis. The result was similar in various sensitivity analyses conducted.

The use of real-world clinical data increases the generalisability of the study’s findings, though given that this data is from a large mental health trust, there is a greater likelihood of severe psychopathology and antipsychotic prescription compared to the general population, which may negatively impact the wider generalisability of findings.

In terms of limitations, a significant hazard ratio for development of Parkinson’s disease was observed for antipsychotic use after adjustment for covariates. With regards to this observed relationship with antipsychotic use, we must first note that the approach taken (two or more mentions of antipsychotics in notes), whilst standard for EHR-based research, may be overly inclusive and potentially include patients in whom antipsychotic medication has been considered but not commenced. Another point for consideration is the propensity for antipsychotics, such as aripiprazole or quetiapine, to be prescribed for treatment-resistant depression. In addition, it is worth noting that some antipsychotics, particularly first-generation antipsychotics, may cause parkinsonism as a side effect. Therefore, there is a possibility that some of the detected PD cases may potentially be misdiagnosed cases of drug-induced parkinsonism though we would not expect this to be the case for clinician-coded diagnostic data from CRIS or HES linkage.

Our findings indicate that there is value in thorough clinical assessment of patients aged over 40 years with a first episode of depression for detection of psychomotor symptoms, given their potential role as a prodromal marker for PD. These study results are generalisable as the CRIS patient record database is a large database of ‘real-world’ patient data from a large mental health trust (Perera et al., 2016). There may be value in including psychomotor retardation alongside other important but often under-recognised prodromal symptoms of Parkinson’s disease, including anosmia and REM sleep behaviour disorder. Whilst present treatments for Parkinson’s disease are focused on symptom reduction, early detection strategies can ensure appropriate referral to tertiary neurology services capable of providing timely care necessary for improving quality of life.

## 5. Conclusion

This study suggests that psychomotor retardation in individuals with depression is associated with higher risk of subsequent Parkinson’s diagnosis and therefore may serve as a prodromal indicator of neurodegeneration. There is a need for prospective longitudinal cohort studies of individuals with depression, incorporating neuroimaging as well as measurement of biochemical and genetic biomarkers (Lotankar et al., 2017) to monitor for evidence of neurodegenerative changes, exploring the potential aetiological link between psychomotor symptoms and Parkinson’s neuropathology.

## Data Availability

The code used for statistical analysis during the current study are available in Supplementary Methods 1.

https://osf.io/2z7gx

## CRediT authorship contribution statement

HM: Writing – original draft, Writing – review & editing, Visualization, Methodology, Formal analysis, Conceptualization.

JB: Writing – review & editing, Visualization, Methodology, Formal analysis, Conceptualization.

EB: Conceptualization.

AF: Methodology, Formal analysis.

ASM: Methodology, Formal analysis.

PS: Supervision.

MZ: Supervision.

AD: Methodology, Supervision.

GL: Methodology, Supervision.

JR: Writing – review & editing, Visualization, Methodology, Formal analysis, Conceptualization, Supervision, Project administration.

## Consent for publication

N/A

## Ethics approval and consent to participate

CRIS has Research Ethics Committee approval as a source of anonymised data for secondary analysis (Oxford REC C, reference 18/SC/0372).

## Declaration of Generative AI and AI-assisted technologies in the writing process

During the preparation of this work, the authors used no generative AI or AI-assisted technologies.

## Acknowledgments

We would like to thank the SLaM CRIS team, especially Amelia Jewell (Research Informatics and Governance Lead) for conducting the search (CRIS project 22-092). This paper represents independent research part-funded by the National Institute for Health Research (NIHR) Biomedical Research Centre at South London and Maudsley NHS Foundation Trust and King’s College London. The views expressed are those of the authors and not necessarily those of the NHS, the NIHR or the Department of Health and Social Care.

