## Supplementary materials for "Psychomotor retardation and risk of Parkinson’s disease in unipolar depression: a retrospective cohort study"

Supplementary Table 1: Definition of variables

| Variable | Source | Definition | Role in analysis |
|---|---|---|---|
| Depression | CRIS structured fields | First ICD-10 coded diagnosis of F32 – Depressive episode or F33 – Recurrent depressive disorder at age 40 years or greater | Inclusion criterion |
| Gender | CRIS structured fields | As defined by patient on entry into healthcare record | Covariate as binary variable |
| Ethnicity | CRIS structured fields | As defined by patient on entry into healthcare record | Covariate (dichotomised into largest group (White) and other) |
| Neurological comorbidity | CRIS free text, identified using the Medical Concept Annotation Tool (MedCAT), an NLP application (NIHR Maudsley BRC, 2025) | A diagnosis of cerebrovascular accident, transient ischaemic attack, multiple sclerosis or epilepsy | Covariate (dichotomised as the presence or absence of any of these comorbidities) |
| Psychiatric comorbidity | CRIS structured fields | Any history of an ICD-10 diagnosis of F00-09 (organic, including symptomatic, mental disorders) or F20-29 (schizophrenia, schizotypal and delusional disorders) in the structured field of the patient records. | Covariate (dichotomised as the presence or absence of any of these comorbidities) |
| Parkinson's disease | CRIS structured fields; CRIS free text; HES | Whichever comes earliest of: (1) a diagnosis of Parkinson's disease coded in | Outcome (earliest of any method of Parkinson's disease diagnosis) |

| | | | |
|---|---|---|---|
| | | CRIS with an ICD-10 code of G20 or F02.3; (2) a diagnosis of Parkinson's disease in the free text of CRIS identified using the Medical Concept Annotation Tool (MedCAT), an NLP application; and (3) a diagnosis of Parkinson's in linked Hospital Episodes Statistics with an ICD-10 code of G20 or F02.3 | |
| Age at depression | CRIS structured fields | Date of birth (to 15th of month) subtracted from date of depression | Covariate |
| Age at psychomotor retardation status | CRIS structured fields; CRIS structured fields | Date of birth (to 15th of month) (in structured field) subtracted from date of first psychomotor status (in free text, analysed by NLP application) | Descriptive statistic |
| Censor date | CRIS structured fields; HES; NHS Spine | Whichever comes earliest of: (1) diagnosis of Parkinson's disease, (2) death (NHS Spine), (3) or 31/03/2023 | Defines follow-up time |
| Electroconvulsive therapy use | CRIS structured field | Presence of an electroconvulsive therapy episode or event between index date and censor. | Descriptive statistic |
| Observation time | CRIS; HES | Index date subtracted from censor date | Defines observation time |

| | | | |
|---|---|---|---|
| Time until Parkinson's disease | CRIS; HES | For patients who develop Parkinson's disease, index date subtracted from first date of Parkinson's diagnosis | Descriptive statistic |
| Age at censor | CRIS; HES | Date of birth (to 15th of month) subtracted from censor date | Descriptive statistic |
| Antipsychotic use | CRIS | At least one mention of antipsychotic medications from index date up to censor date (in free text, analysed by NLP application) | Covariate |
| Death | NHS Spine | Date of death | Competing event |

CRIS – Clinical Records Interactive Search; HES – Hospital Episode Statistics; NHS – National Health Service; NLP - Natural Language Processing

Supplementary Table 2: STROBE checklist

| | Item No | Recommendation | Page No |
|---|---|---|---|
| **Title and abstract** | 1 | (*a*) Indicate the study's design with a commonly used term in the title or the abstract | 1 |
| | | (*b*) Provide in the abstract an informative and balanced summary of what was done and what was found | 2 |
| **Introduction** | | | |
| Background /rationale | 2 | Explain the scientific background and rationale for the investigation being reported | 2-3 |
| Objectives | 3 | State specific objectives, including any prespecified hypotheses | 3 |
| **Methods** | | | |
| Study design | 4 | Present key elements of study design early in the paper | 3-4 |
| Setting | 5 | Describe the setting, locations, and relevant dates, including periods of recruitment, exposure, follow-up, and data collection | 3-4 |
| Participants | 6 | (*a*) Give the eligibility criteria, and the sources and methods of selection of participants. Describe methods of follow-up | 3-4 |

| | | | |
|---|---|---|---|
| | | (*b*) For matched studies, give matching criteria and number of exposed and unexposed | 5-6 |
| Variables | 7 | Clearly define all outcomes, exposures, predictors, potential confounders, and effect modifiers. Give diagnostic criteria, if applicable | 4-5 |
| Data sources/ measureme nt | 8* | For each variable of interest, give sources of data and details of methods of assessment (measurement). Describe comparability of assessment methods if there is more than one group | *S1* |
| Bias | 9 | Describe any efforts to address potential sources of bias | 5 |
| Study size | 10 | Explain how the study size was arrived at | 5-6 |
| Quantitative variables | 11 | Explain how quantitative variables were handled in the analyses. If applicable, describe which groupings were chosen and why | 5 |
| Statistical methods | 12 | (*a*) Describe all statistical methods, including those used to control for confounding | 5 |
| | | (*b*) Describe any methods used to examine subgroups and interactions | 5 |
| | | (*c*) Explain how missing data were addressed | 4 |
| | | (*d*) If applicable, explain how loss to follow-up was addressed | NA |
| | | (*e*) Describe any sensitivity analyses | 5 |
| **Results** | | | |
| Participants | 13* | (a) Report numbers of individuals at each stage of study—eg numbers potentially eligible, examined for eligibility, confirmed eligible, included in the study, completing follow-up, and analysed | 5 |
| | | (b) Give reasons for non-participation at each stage | NA |
| | | (c) Consider use of a flow diagram | NA |
| Descriptive data | 14* | (a) Give characteristics of study participants (eg demographic, clinical, social) and information on exposures and potential confounders | 6 |
| | | (b) Indicate number of participants with missing data for each variable of interest | 6 |
| | | (c) Summarise follow-up time (eg, average and total amount) | 6 |
| Outcome data | 15* | Report numbers of outcome events or summary measures over time | 7 |
| Main results | 16 | (*a*) Give unadjusted estimates and, if applicable, confounder-adjusted estimates and their precision (eg, 95% confidence interval). Make clear which confounders were adjusted for and why they were included | 7 |
| | | (*b*) Report category boundaries when continuous variables were categorized | NR |
| | | (*c*) If relevant, consider translating estimates of relative risk into absolute risk for a meaningful time period | 7 |
| Other analyses | 17 | Report other analyses done—eg analyses of subgroups and interactions, and sensitivity analyses | 7 |
| **Discussion** | | | |
| Key results | 18 | Summarise key results with reference to study objectives | 7 |
| Limitations | 19 | Discuss limitations of the study, taking into account sources of potential bias or imprecision. Discuss both direction and magnitude of any potential bias | 9 |
| Interpretatio n | 20 | Give a cautious overall interpretation of results considering objectives, limitations, multiplicity of analyses, results from similar studies, and other relevant evidence | 9 |
| Generalisab ility | 21 | Discuss the generalisability (external validity) of the study results | 9-10 |

| Other information | | | |
|---|---|---|---|
| Funding | 22 | Give the source of funding and the role of the funders for the present study and, if applicable, for the original study on which the present article is based | 11 |

Supplementary Table 3: Backwards logistic mixed-effects model with psychomotor retardation exposure status as the dependent variable

| Independent variable | OR | 95% CI (lower – upper) | p-value |
|---|---|---|---|
| **Intercept** | 0.061 | 0.034 – 0.11 | <0.001 |
| **Linear component of time** | $1.86x10^{29}$ | $1.01\times10^{25}$ – $3.44\times10^{34}$ | <0.001 |
| **Quadratic component of time** | $7.65x10^{5}$ | 56.68 – $1.03\times10^{10}$ | 0.005 |
| **Parkinson's disease diagnosis** | 2.493 | 1.21 – 5.16 | 0.014 |
| **Age at depression diagnosis** | 1.09 | 1.06 – 1.12 | <0.001 |
| **Age at censor** | 0.94 | 0.91 – 0.96 | <0.001 |
| **Gender** | 0.92 | 0.75 – 1.14 | 0.473 |
| **Ethnicity** | 0.28 | 0.22 – 0.34 | <0.001 |
| **Antipsychotic use** | 3.43 | 2.78 – 4.23 | <0.001 |
| **Neurological comorbidities** | 0.39 | 0.21 – 0.74 | 0.004 |
| **Psychiatric comorbidities** | 0.81 | 0.52 – 1.25 | 0.338 |
| **Interaction between Parkinson's status and linear time component** | $4.53x10^{17}$ | $1.23\times10^{-21}$ – $1.67\times10^{56}$ | 0.370 |
| **Interaction between Parkinson's status and quadratic time component** | $6.08\times10^{14}$ | $1.12\times10^{-13}$ – $3.31\times10^{42}$ | 0.296 |

Supplementary methods 1: R analysis code

Available at https://osf.io/2z7gx
